# Assessing the utility of lymphocyte count to diagnose COVID-19

**DOI:** 10.1101/2021.03.17.21252922

**Authors:** Michael Fralick, Orly Bogler, Daniel Tamming, Lauren Lapointe-Shaw, Janice Kwan, Terence Tang, Shail Rawal, Jessica Liu, Farad Razak, Amol A Verma

## Abstract

**Background:** COronaVirus Disease 2019 (COVID-19) can be challenging to diagnose, because symptoms are non-specific, clinical presentations are heterogeneous, and false negative tests can occur. Our objective was to assess the utility of lymphocyte count to differentiate COVID-19 from influenza or community-acquired pneumonia (CAP).

**Methods:** We conducted a cohort study of adults hospitalized with COVID-19 or another respiratory infection (i.e., influenza, CAP) at seven hospitals in Ontario, Canada.The first available lymphocyte count during the hospitalization was used. Standard test characteristics for lymphocyte count (×10^9^/L) were calculated (i.e., sensitivity, specificity, area under the receiver operating curve [AUC]). All analyses were conducting using R.

**Results:** There were 869 hospitalizations for COVID-19, 669 for influenza, and 3009 for CAP. The mean age across the three groups was 67 and patients with pneumonia were older than those with influenza or COVID19, and approximately 46% were woman. The median lymphocyte count was nearly identical for the three groups of patients: 1.0 ×10^9^/L (interquartile range [IQR]:0.7,2.0) for COVID-19, 0.9 ×10^9^/L (IQR 0.6,1.0) for influenza, and 1.0 ×10^9^/L (IQR 0.6,2.0) for CAP. At a lymphocyte threshold of less than 2.0 ×10^9^/L, the sensitivity was 87% and the specificity was approximately 10%. As the lymphocyte threshold increased, the sensitivity of diagnosing COVID-19 increased while the specificity decreased. The AUC for lymphocyte count was approximately 50%.

**Interpretation:** Lymphocyte count has poor diagnostic discrimination to differentiate between COVID-19 and other respiratory illnesses. The lymphopenia we consistently observed across the three illnesses in our study may reflect a non-specific sign of illness severity. However, lymphocyte count above 2.0 ×10^9^/L may be useful in ruling out COVID-19 (sensitivity = 87%).

## Introduction

COronaVirus Disease 2019 (COVID-19) can be challenging to diagnose, because symptoms are non-specific, clinical presentations are heterogeneous, and false negative tests can occur.^1^ While most patients hospitalized with COVID-19 will have lymphopenia, it is unknown if lymphopenia can differentiate between COVID-19 and other respiratory infections.^2^ Our objective was to assess the utility of lymphocyte count to differentiate COVID-19 from influenza or community-acquired pneumonia (CAP).

## Methods

We conducted cohort study of adults hospitalized with COVID-19 or another respiratory infection (i.e., influenza, CAP) at seven hospitals in Ontario, Canada.^3^ The influenza cohort and CAP cohort spanned September to April, 2020, and the COVID-19 cohort spanned January to June, 2020 (most recent available data). CAP was identified using a previously developed definition combining a diagnosis of CAP with receipt of antibiotic that is typically used for CAP (in-press). Influenza was defined using ICD10 diagnostic codes (positive-predictive-value > 90%).^4^ Patients with COVID-19 who also met the criteria for CAP or influenza were assigned to the COVID-19 group. The first available lymphocyte count during the hospitalization was used. Standard test characteristics for lymphocyte count (×10^9^/L) were calculated (i.e., sensitivity, specificity, area under the receiver operating curve [AUC]). All analyses were conducting using R.

## Results

There were 869 hospitalizations for COVID-19, 669 for influenza, and 3009 for CAP. The mean age across the three groups was 67 and patients with pneumonia were older than those with influenza or COVID19, and approximately 46% were woman. The median lymphocyte count was nearly identical for the three groups of patients: 1.0 ×10^9^/L (interquartile range [IQR]:0.7,2.0) for COVID-19, 0.9 ×10^9^/L (IQR 0.6,1.0) for influenza, and 1.0 ×10^9^/L (IQR 0.6,2.0) for CAP. At a lymphocyte threshold of 1.0 the sensitivity and specificity for diagnosing COVID was less than 50% (Table 1). At higher lymphocyte thresholds the sensitivity increased and the specificity decreased (Table 1). For example, at a threshold of 2.0 the sensitivity and specificity was 87% and approximately 10%, respectively. The AUC for lymphocyte count was 54% when patients with influenza were the comparator and 51% when patients with CAP were the comparator (Table 1, Figure 1).

**Table 1.** Baseline characteristics and test characteristics of lymphocyte count

|  | COVID-19<br>(N=869) | Influenza<br>(N=669) | CAP<br>(N=3009) |
| --- | --- | --- | --- |
| Age, Years, mean (SD) | 65.7 (17.4) | 66.1 (18.1) | 69.7 (17.8) |
| Female, Sex (%) | 365 (42) | 334 (50) | 1315 (44) |
| Lymphocyte count |  |  |  |
| Median (IQR) | 1.0 (0.7, 2.0) | 0.9 (0.6, 1.0) | 1.0 (0.6, 2.0) |
| < 1.0 (%) | 421 (48) | 353 (53) | 1371 (46) |
| < 1.5 (%) | 646 (74) | 517 (77) | 2151 (71) |
| < 2.0 (%) | 756 (87) | 600 (90) | 2559 (85) |
| **Test characteristics for lymphocyte count for the diagnosis of COVID-19** |  |  |  |
| Area under the curve (95% CI) | 0.54 (0.51, 0.57) [flu comp] 0.51 (0.49, 0.53) [CAP comp] |  |  |
| Sensitivity |  |  |  |
| Lymphocyte < 1.0 (95% CI) | 0.48 (0.45, 0.52) |  |  |
| Lymphocyte < 1.5 (95% CI) | 0.74 (0.71, 0.77) |  |  |
| Lymphocyte < 2.0 (95% CI) | 0.87 (0.85, 0.89) |  |  |
| Specificity |  |  |  |
| Lymphocyte < 1.0 (95% CI) | 0.47 (0.43, 0.51) [flu comp] 0.54 (0.53, 0.56) [CAP comp] |  |  |
| Lymphocyte < 1.5 (95% CI) | 0.23 (0.20, 0.26) [flu comp] 0.29 (0.27, 0.30) [CAP comp] |  |  |
| Lymphocyte < 2.0 (95% CI) | 0.10 (0.08, 0.13) [flu comp] 0.15 (0.14, 0.16) [CAP comp] |  |  |

Legend: COVID-19 COroNaVirus Disease 2019, flu = influenza, CAP = community-acquired pneumonia, IQR = interquartile range, SD = standard deviation. Comp = comparator. Note there is only one sensitivity value and two specificity values because the former is calculated as the true positive / all patients with COVID (i.e., true positive plus false negative) while the specificity is calculated as the true negative / all patients who did not have COVID (i.e., they could have had pneumonia or influenza defined as true negative plus false positive)

**Figure 1.**
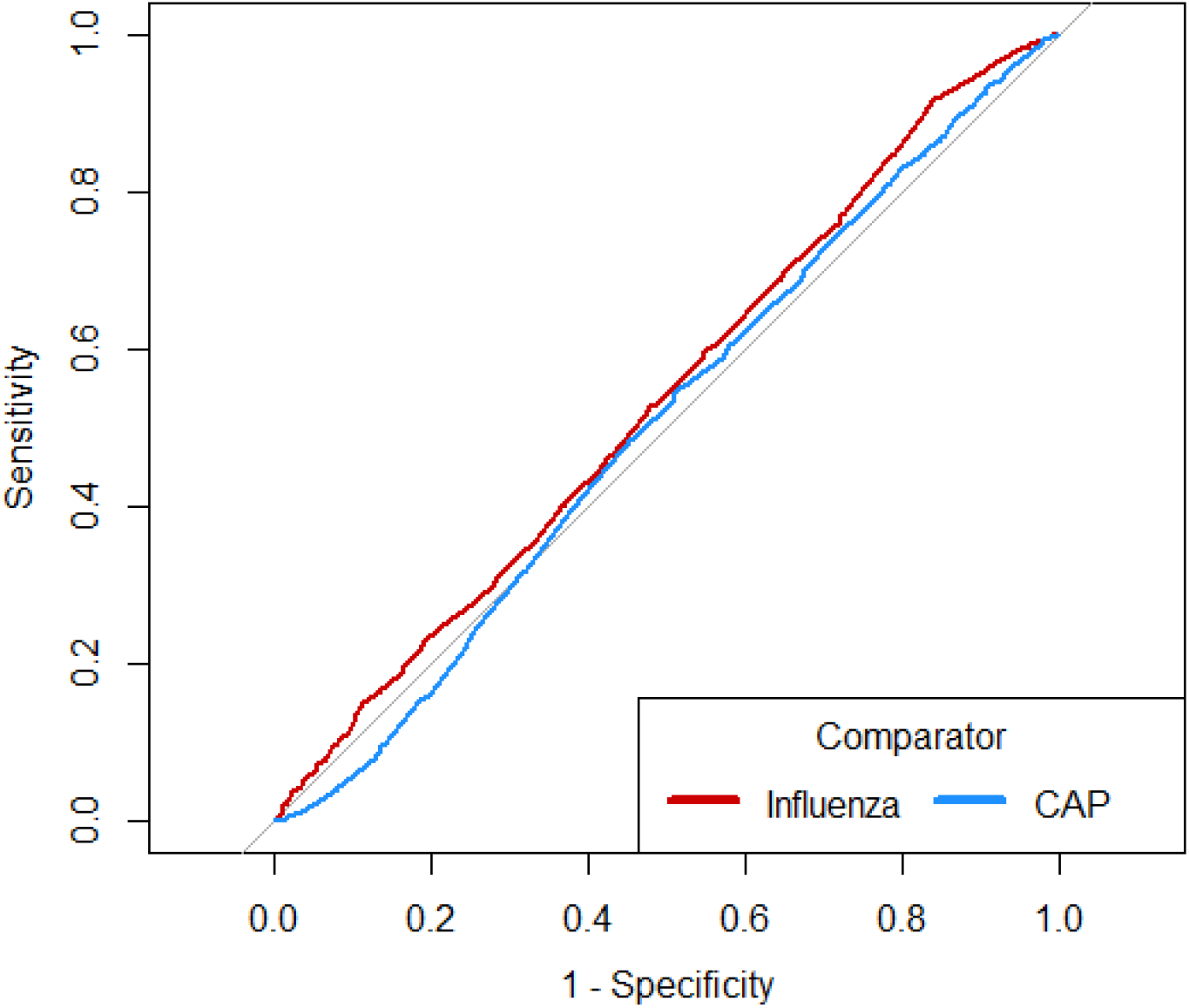
Receiver operating curve for lymphocyte count and diagnosis of COVID-19 1. Legend: CAP = community acquired pneumonia

## Discussion

Among hospitalized adults, the lymphocyte count was similar for patients with COVID-19, influenza, and CAP. The AUC was close to 0.5, indicating that lymphocyte count has similar accuracy to a coin-toss in determining whether the patient has COVID-19 or another respiratory illness. Thus, lymphocyte count alone is unlikely to be useful to differentiate between COVID-19 and other respiratory illnesses. While lymphopenia is non-specific and thus does not help to rule in COVID-19, the absence of a lymphocyte count above 2.0 may help to rule out COVID-19 (sensitivity of 87%). It is unknown if these findings also apply to patients with mild disease who are not hospitalized. Another important limitation of our study is that we lacked data on chronic medical conditions pre-dating the hospitalization known to cause lymphopenia (e.g., hematologic malignancies, autoimmune disorders), however, we expect these conditions to be rare and not differential across the three respiratory illnesses. The lymphopenia we consistently observed across the three illnesses in our study may reflect a non-specific sign of illness severity, with limited diagnostic utility in identifying underlying etiologies.^5^

## Data Availability

Not available.

## Notes

**Obtained funding:** This study was funded by Sinai Health and the Canadian Institutes of Health Research (Grant Number VR4 - 172743). The development of the GEMINI data platform has been supported with funding from the Canadian Cancer Society, the Canadian Frailty Network, the Canadian Institutes of Health Research, the Canadian Medical Protective Agency, Green Shield Canada Foundation, the Natural Sciences and Engineering Research Council of Canada, Ontario Health, the St. Michael’s Hospital Association Innovation Fund, the University of Toronto Department of Medicine, and in-kind support from partner hospitals and Vector Institute. F.R. is supported by an award from the Mak Pak Chiu and Mak-Soo Lai Hing Chair in General Internal Medicine, University of Toronto.

**Conflicts of interest:** Amol Verma and Fahad Razak are employees of Ontario Health, outside of this project. Mike Fralick is a consultant for a start-up company based at the Massachusetts Institute of Technology (MIT) developing a CRISPR based diagnostic test for COVID19.

### Competing Interest Statement

Amol Verma and Fahad Razak are employees of Ontario Health, outside of this project. Mike Fralick is a consultant for a start-up company based at the Massachusetts Institute of Technology (MIT) developing a CRISPR based diagnostic test for COVID19.

### Funding Statement

This study was funded by Sinai Health and the Canadian Institutes of Health Research (Grant Number VR4-172743).  The development of the GEMINI data platform has been supported with funding from the Canadian Cancer Society, the Canadian Frailty Network, the Canadian Institutes of Health Research, the Canadian Medical Protective Agency, Green Shield Canada Foundation, the Natural Sciences and Engineering Research Council of Canada, Ontario Health, the St. Michaels Hospital Association Innovation Fund, the University of Toronto Department of Medicine, and in-kind support from partner hospitals and Vector Institute. F.R. is supported by an award from the Mak Pak Chiu and Mak-Soo Lai Hing Chair in General Internal Medicine, University of Toronto.

### Author Declarations

Approved by St Michael's Hospital REB

